# Women’s Empowerment and Quality Antenatal Care in Ghana: Analysis of the SWPER Global Index in the Ghana Demographic Health Survey

**DOI:** 10.1101/2025.02.25.25322866

**Authors:** Barnabas Bessing, Munawar Harun Koray, Linus Baatiema

**Affiliations:** Department of Public Health, Ghana Health Service, Upper West Regional Health Directorate, Wa, Ghana; Southern Medical University, Guangzhou, China; Centre for Environment, Migration and International Relations, Simon Diedong Dombo The University of Business and Integrated Development Studies, Wa, Ghana; School of Public Health, Faculty of Public Health, University of Port Harcourt, Port Harcourt, Nigeria

**Keywords:** Women’s empowerment, antenatal care, Sustainable Development Goals, SWPER index, Ghana, maternal health, demographic health survey

## Abstract

**Background:** Women’s empowerment is a key determinant of maternal healthcare utilization, influencing antenatal care (ANC) quality in low- and middle-income countries (LMICs). While ANC is crucial for maternal and neonatal health outcomes, disparities persist in service uptake and quality in Ghana. This study examines the association between women’s empowerment, measured using the Survey-based Women’s emPowERment (SWPER) index, and the receipt of quality ANC in Ghana.

**Methods:** A cross-sectional analysis was conducted using the 2022 Ghana Demographic and Health Survey (GDHS) data. The study included 8,715 women aged 15–49 who attended ANC during pregnancy. Quality ANC received four essential services: blood pressure monitoring, urine testing, iron supplementation, and blood sample collection. Women’s empowerment was assessed across three SWPER domains: attitude toward violence, decision-making, and social independence, categorised into low, medium, and high levels. Logistic regression models examined the relationship between empowerment and quality ANC while adjusting for demographic and socioeconomic variables.

**Results:** The study found that 95% of women received quality ANC services despite significant disparities across demographic and socioeconomic groups. Higher levels of women’s empowerment were positively associated with quality ANC. Specifically, women with high empowerment in attitude toward violence (AOR: 1.92; 95% CI: 1.53 - 3.10), social independence (AOR: 1.82; 95% CI: 1.69 - 1.99), and decision-making (AOR: 1.31; 95% CI: 1.09 - 1.58) were significantly more likely to receive quality ANC than their low-empowerment counterparts. Other significant predictors of quality ANC included higher education, wealth index, older maternal age, urban residence, and media exposure. Women in rural areas and those in the Northern, Northeast, and Savannah regions were significantly less likely to receive quality ANC.

**Conclusion:** Women’s empowerment is a key driver of quality ANC utilisation in Ghana. Strengthening empowerment initiatives such as increasing educational opportunities, enhancing economic autonomy, and promoting decision-making power can improve maternal health outcomes. Addressing regional disparities and integrating empowerment strategies into maternal health policies can enhance ANC quality and reduce maternal and neonatal mortality in Ghana.

---

Antenatal care (ANC) is a critical component of maternal healthcare, ensuring positive pregnancy outcomes and reducing maternal and neonatal morbidity and mortality. Quality ANC encompasses the frequency of visits and the comprehensiveness of care, including essential services such as blood pressure monitoring, urine testing, iron supplementation, and blood sample collection.[1, 2]. The World Health Organization (WHO) initially recommended a minimum of four ANC visits. Still, it revised its guidelines in 2016 to advocate for at least eight contacts, emphasising a holistic and continuous approach to maternal care. [1].

Despite global standards, significant gaps persist in providing quality ANC, particularly in low- and middle-income countries (LMICs) such as Ghana. Previous research in developing countries has shown that factors such as educational attainment, economic status, and timely ANC initiation are critical determinants of ANC quality.[3-6]. In Ghana, regional disparities, socioeconomic barriers, and cultural factors hinder equitable access to quality maternal healthcare services.[7-11].

Women’s empowerment has emerged as a significant determinant of maternal healthcare utilisation, particularly in resource-constrained settings. Empowered women who possess greater decision-making autonomy, social independence, and resistance to gender-based violence are more likely to seek higher-quality ANC services.[2, 12-15]. The Survey-based Women’s emPowERment (SWPER) global index provides a validated measure to assess women’s empowerment across three key domains: attitude toward violence, decision-making, and social independence, which have been shown to influence maternal health-seeking behaviour.[13].

The 2022 Ghana Demographic and Health Survey (GDHS) offers a comprehensive dataset to examine the relationship between women’s empowerment and quality ANC in Ghana. While previous studies have established the importance of women’s empowerment in maternal healthcare utilisation, a gap exists in understanding how empowerment affects the quality of ANC received by Ghanaian women. This study seeks to bridge this gap by analysing the association between women’s empowerment using SWPER domains (decision-making, social independence, and attitudes toward violence) and quality ANC while accounting for sociodemographic factors and regional disparities.

Given Ghana’s persistent urban-rural divide and regional inequalities, understanding the factors influencing quality ANC uptake is crucial for developing targeted maternal health interventions. This study aims to contribute evidence-based insights to inform policies that enhance maternal healthcare access, empower women, and improve pregnancy outcomes across diverse socioeconomic contexts in Ghana.

## METHODS

### Study design and data source

This research employed a cross-sectional design using the latest (2022) Ghana Demographic and Health Survey (GDHS). The Demographic and Health Survey (DHS), a nationally representative survey, is carried out every five years in over 85 low- and middle-income countries globally [16] . Participants for the survey are chosen using a multistage sampling technique. This approach guarantees that the sample is representative enough of the populations at national, regional (e.g., departments, states) and place of residence (urban-rural) levels, including household and community characteristics. The DHS program collects data from 5,000 to 32,000 participants using a uniform questionnaire. This questionnaire covers a range of variables, including demographic and socioeconomic factors, family planning, infant and child mortality, reproductive and general women’s health, HIV/AIDS, and women empowerment, amongst others (see DHS website for details) and in literature [16, 17].

### Sample size

From the GDHS 2022, 18,450 households were selected in 618 clusters, where 15,014 women aged 15–49 and 7,044 men aged 15–59 were interviewed in one of every two households selected during the GDHS. In this study, we included 58% (8,715) of the women surveyed during the GDHS because they indicated they attended ANC during their pregnancy at the time of the data collection.

### Study measures

#### Outcome variable

Our primary outcome variable was “Quality ANC” adopted from a previous study [2] and is defined as the proportion of mothers who have received all four essential services such as 1) blood pressure monitoring, 2) urine testing, 3) supply of iron supplement to be taken at home, and 4) collection of a blood sample for testing during any ANC visit. Pregnant women who received all four or more services during ANC visits were classified as having received quality ANC (coded as 1). In comparison, those who did not receive all four services were categorised otherwise (coded as 0).

#### Independent variables

The primary independent variable for this study is the level of women empowerment according to the SWPER global index, which was developed using the DHS for 62 lower-middle-income countries [18, 19]. The SWPER consist of three key domains based on 15 questions from the DHS women questionnaire: 1) attitude towards violence, 2) social independence, and 3) decision making. The attitude towards violence domain evaluates a woman’s belief on whether it is ever acceptable for a husband to beat his wife in certain situations consisting of five questions. The social independence domain examines a woman’s access to information, educational attainment, age at marriage and first childbirth, and the age and education differences between her partners. It consists of six questions. The decision-making domain assesses who makes household decisions and the woman’s employment status consisting of four questions. Based on these, SWPER classified women’s empowerment levels into low, medium, and high, as described by Ewerling et al [18, 19].

#### Demographic and socioeconomic characteristics

Based on a meta-analysis of significant predictors of ANC in published and unpublished DHS studies from 2002-2022 [20], we included the following covariates in our analysis: age, religion, educational level, wealth index, frequency of listening to radio, frequency of watching television, frequency of reading newspaper/magazine, place of residence, and region.

### Data Analysis

Descriptive statistics were presented as frequencies with percentages for categorical variables. Pearson’s chi-square test was employed to determine if there is a significant difference in outcome quality ANC (Yes/No) amongst women empowerment, demographic and socioeconomic variables.

To evaluate the associations with our dichotomous outcome (quality ANC), we compared two methods of logistic regression modelling to obtain crude odds ratios (COR) and adjusted odds ratios (AOR). We first fitted a bivariate logistic regression to determine if the women empowerment levels, demographic, and socioeconomic variables were associated with quality ANC. A final single multivariable logistic regression model was fitted that, at the outset, included all variables which were significant (p-value <0.05) in the bivariate regression analyses and compared that with a stepwise backward regression elimination multivariable model using the Akaike Information Criterion (AIC)[21]. We selected the backward elimination model using the AIC as our final model because it minimizes overfitting and was robust, consistent, and reliable in its variable selection.

Given the binary nature of the outcome (quality ANC: Yes/No), we opted for logistic regression to estimate the association. Poisson regression was considered but not implemented due to the relatively low prevalence of the non-receipt of quality ANC which could bias the standard errors. Additionally, since data is nested within the households and regions, a multilevel logistic regression model was tested to adjust for intra-cluster correlation. Still, likelihood-ratio testing (p=0.23) indicated that a single-level logistic model was sufficient. Missing data were handled using the maximum likelihood estimation with missing at random assumption.

Variance inflation factor (VIF) was used to test for multicollinearity, and there was no evidence of multicollinearity in the models since all predictor variables had VIF < 2.72. Education level and wealth index showed a moderate correlation (VIF=2.5), but separate inclusion was justified based on theoretical consideration. The final model goodness of fit was examined using Hosmer-Lemeshow goodness-of-fit and the goodness of fit was good (Hosmer-Lemeshow chi-square (6) = 7.86; p-value = 0.28). Model discrimination was assessed using the area under the receiver operating characteristics curve (AUC-ROC = 0.83), indicating strong predictive ability. The Survey-Weighted Nagelkerke’s R^2^ value was 0.32, suggesting that women empowerment and socioeconomic variables explained 32% of the variance in quality ANC receipt. We used the “svy” STATA command to take into consideration the complex survey design of the DHS program dataset.

Sensitivity analyses were conducted to test the robustness of our results. First, logistics regression models were run with and without survey weights, showing no substantial difference in estimates. Then stratified analyses by urban/rural residence confirmed that empowerment effects were stronger in urban settings. To assess potential self-selection bias, we estimated a Heckman two-step selection model to test if unobserved factors influencing empowerment also systematically impact ANC quality using the Inverse Mills ratio (IMR). All analyses were done using STATA version 18.0.

This study utilised the DHS database, encompassing a global survey conducted over five years. Inner City Fund (ICF) International authorised access to the data after registering the topic and submitting a request on their website. Comprehensive details on the methodology and ethical considerations can be found on the DHS website. According to the ICF International Institutional Review Board, individual consent was unnecessary as the data were secondary and publicly available. The study obtained all required approvals from the DHS Program, and data privacy was strictly upheld, including no personal identifiers during processing and analysis.

## RESULTS

### Participant characteristics

In **Table 1**, the majority (68%0 of the participants were ≥30 years old, 72% were Christians, 65% had primary and secondary education, 42% were in the rich wealth index, and 52% were urban dwellers, with 60% of them able to watch television more than once a week. Only four of the sixteen regions consisting of Ashanti (17%), Greater Accra (14%), Northern (11%) and Central (10%) contributed about 52% of the total participants.

**Table 1:** Association between quality ANC and covariates.

| Characteristics | Weighted<br>Frequency<br>(n = 8715) | Per cent<br>(%) | Quality ANC | | $\chi^2$ |
| --- | --- | --- | --- | --- | --- |
|  |  |  | No (%) | Yes (%) |  |
| Age categories |  |  |  |  |  |
| 15-19 | 207 | 2.38 | 22 (11.60) | 170 (88.40) | 11.41* |
| 20-24 | 1075 | 12.33 | 83 (8.29) | 918 (91.71) |  |
| 25-29 | 1554 | 17.83 | 90 (6.22) | 1,357 (93.78) |  |
| 30-34 | 1818 | 20.86 | 89 (5.23) | 1,604 (94.77) |  |
| 35-39 | 1753 | 20.11 | 74 (4.52) | 1,557 (95.48) |  |
| 40-44 | 1310 | 15.03 | 40 (3.26) | 1,180 (96.74) |  |
| 45-49 | 998 | 11.46 | 10 (1.05) | 919 (98.95) |  |
| Religion |  |  |  |  |  |
| Christianity | 6,244 | 71.65 | 225 (3.87) | 5,586 (96.13) | 34.64* |
| Islam | 2,040 | 23.41 | 119 (6.24) | 1,780 (93.76) |  |
| Other | 248 | 2.85 | 46 (20.06) | 185 (79.94) |  |
| No religion | 183 | 2.1 | 17 (10.05) | 153 (89.95) |  |
| Educational level |  |  |  |  |  |
| No education | 2,131 | 24.45 | 170 (8.58) | 1,813 (91.42) | 18.19* |
| Primary | 1,306 | 14.98 | 75 (6.18) | 1,140 (93.82) |  |
| Secondary | 4,437 | 50.91 | 139 (3.36) | 3,990 (96.64) |  |
| Tertiary | 841 | 9.66 | 23 (2.91) | 760 (97.09) |  |
| Wealth index |  |  |  |  |  |
| Poorest | 1,775 | 20.37 | 167 (10.08) | 1,486 (89.92) | 18.47* |
| Poorer | 1,599 | 18.35 | 86 (5.76) | 1,403 (94.24) |  |
| Middle | 1,640 | 18.82 | 62 (4.04) | 1,465 (95.96) |  |
| Richer | 1,832 | 21.02 | 49 (2.89) | 1,656 (97.11) |  |
| Richest | 1,869 | 21.44 | 44 (2.51) | 1,696 (97.49) |  |
| Frequency of reading newspaper |  |  |  |  |  |
| Not at all | 8,004 | 91.85 | 383 (5.14) | 7,067 (94.86) | 0.85 |
| < once a week | 493 | 5.65 | 19 (4.04) | 440 (95.96) |  |
| ≥ once a week | 218 | 2.5 | 5 (2.70) | 197 (97.30) |  |
| Frequency of listening to the radio |  |  |  |  |  |
| Not at all | 2,887 | 33.12 | 221 (8.21) | 2,466 (91.79) | 29.06* |
| < once a week | 1,994 | 22.88 | 75 (4.01) | 1,781 (95.99) |  |
| ≥ once a week | 3,834 | 44 | 112 (3.13) | 3,457 (96.87) |  |
| Frequency of watching television |  |  |  |  |  |
| Not at all | 2,279 | 26.15 | 188 (8.84) | 1,933 (91.16) | 31.95* |
| < once a week | 1,176 | 13.5 | 35 (3.23) | 1,060 (96.77) |  |
| ≥ once a week | 5,260 | 60.35 | 184 (3.76) | 4,711 (96.24) |  |

**Table 1 continued:** Association between quality ANC and covariates
| Characteristics | Weighted<br>Frequency<br>(n = 8715) | Per cent<br>(%) | Quality ANC | | $\chi^2$ | |
| --- | --- | --- | --- | --- | --- | --- |
|  |  |  | No (%) | Yes (%) |  |  |
| **Place of residence** |  |  |  |  |  |  |
| Urban | 4,500 | 51.63 | 137 (3.28) | 4,051 (96.72) | 27.01* |  |
| Rural | 4,215 | 48.37 | 270 (6.88) | 3,653 (93.12) |  |  |
| **Region** |  |  |  |  |  |  |
| Western | 523 | 6 | 19 (4.00) | 467 (96.00) | 12.06* |  |
| Central | 863 | 9.91 | 27 (3.39) | 776 (96.61) |  |  |
| Greater Accra | 1211 | 13.9 | 27 (2.38) | 1,100 (97.62) |  |  |
| Volta | 400 | 4.59 | 6 (1.52) | 367 (98.48) |  |  |
| Eastern | 677 | 7.77 | 16 (2.46) | 615 (97.54) |  |  |
| Ashanti | 1508 | 17.31 | 47 (3.35) | 1,357 (96.65) |  |  |
| Western North | 242 | 2.78 | 17 (7.75) | 208 (92.25) |  |  |
| Ahafo | 196 | 2.25 | 4 (2.18) | 179 (97.82) |  |  |
| Bono | 301 | 3.46 | 6 (2.06) | 275 (97.94) |  |  |
| Bono East | 398 | 4.56 | 25 (6.87) | 345 (93.13) |  |  |
| Oti | 264 | 3.03 | 8 (3.24) | 238 (96.76) |  |  |
| Northern | 930 | 10.68 | 116 (13.35) | 750 (86.65) |  |  |
| Savannah | 229 | 2.63 | 29 (13.74) | 184 (86.26) |  |  |
| North East | 244 | 2.8 | 26 (11.65) | 201 (88.35) |  |  |
| Upper East | 452 | 5.18 | 30 (7.08) | 391 (92.92) |  |  |
| Upper West | 276 | 3.16 | 3 (1.36) | 253 (98.64) |  |  |
| ANC: Antenatal Care |  |  |  |  |  |  |
| * = p – value < 0.001 |  |  |  |  |  |  |

### Trends in quality antenatal care

A significant proportion (95%) of pregnant women in this study indicated they had received quality ANC. In **Table 1**, there were significant trends in quality antennal care among participants’ characteristics. For instance, significant trends were observed in quality antenatal care in age categories (***χ***2 = 11.41, p < 0.001), religion (***χ***2 = 34.64, p < 0.001), educational level (***χ***2 = 18.19, p < 0.001), wealth index (***χ***2 = 18.47, p < 0.001), frequency of radio listening per week (***χ***2 = 29.06, p < 0.001), frequency of television watching per week (***χ***2 = 31.95, p < 0.001), and place of residence (***χ***2 = 27.01, p < 0.001).

Similarly, in **Table 2**, significant trends were observed in quality antenatal care amongst the women empowerment index domains. There were significant trends in quality antenatal care in women empowerment levels in the ‘attitude toward violence’ domain ((***χ***2 = 32.36, p < 0.001), ‘decision making’ domain (***χ***2 = 12.88, p < 0.001) and the ‘self-independence’ domain (***χ***2 = 19.58, p < 0.001)

**Table 2:** Association between women empowerment variables levels and quality ANC.

| SWPER domains and level of empowerment | Quality of ANC (Weighted) | | $\chi^2$ |
| --- | --- | --- | --- |
|  | No (%) | Yes (%) |  |
| **Attitude toward violence** |  |  |  |
| Low empowerment | 72 (10.69) | 601 (89.31) | 32.36* |
| Medium empowerment | 86 (8.5) | 931 (91.5) |  |
| High empowerment | 249 (3.87) | 6172 (96.13) |  |
| **Decision making** |  |  |  |
| Low empowerment | 82 (8.51) | 881 (91.49) | 12.88* |
| Medium empowerment | 151 (5.73) | 2,489 (94.27) |  |
| High empowerment | 174 (3.86) | 4,335 (96.14) |  |
| **Social independence** |  |  |  |
| Low empowerment | 110 (6.81) | 1,502 (93.19) | 19.58* |
| Medium empowerment | 182 (6.46) | 2,630 (93.54) |  |
| High empowerment | 116 (3.13) | 3,573 (96.87) |  |
| SWPER: survey-based women's empowerment index; ANC: Antenatal Care |  |  |  |
| * = p – value < 0.001 |  |  |  |

### Demographic, socioeconomic, and women empowerment variables association with quality antenatal care

The bivariate logistic regression analyses in **Table 3** showed that increasing age ≥ 25 years, religion, educational level, wealth index, frequency of radio listening per week, frequency of television watching per week, place of residence and levels of women empowerment were associated with increased likelihood of receiving quality antenatal care. For instance, compared with pregnant women aged 15-19, those in 25-29 (COR: 1.98; 95% CI: 1.08-3.62), 30-34 (COR: 2.38; 95% CI: 1.30-4.33), 35-39 (COR: 2.77; 95% CI: 1.53-5.03), 40-44 (COR: 3.90; 95% CI: 1.94-7.84), 45-49 (COR: 12.32; 95% CI: 5.53-27.45) age categories were more likely to receive quality antenatal care.

**Table 3:** Logistic regression model for ANC determinants.

| **Variables** | **COR (95%CI)** | **AOR (95%CI)** |
| --- | --- | --- |
| **Age categories** |  |  |
| 15-19 | **Ref (1.00)** | **Ref (1.00)** |
| 20-24 | 1.45 (0.79 - 2.68) | - |
| 25-29 | **1.98 (1.08 - 3.62)** | - |
| 30-34 | **2.38 (1.30 - 4.33)** | - |
| 35-39 | **2.77 (1.53 - 5.03)** | **1.30 (1.02 - 1.66)** |
| 40-44 | **3.90 (1.94 - 7.84)** | **1.96 (1.44 - 2.68)** |
| 45-49 | **12.32 (5.53- 27.45)** | **4.55 (2.76 - 7.49)** |
| **Religion** |  |  |
| Christianity | **Ref (1.00)** | **Ref (1.00)** |
| Islam | **0.61 (0.45 - 0.81)** | - |
| Other religion | **0.16 (0.11 - 0.24)** | **0.39 (0.27 - 0.57)** |
| No religion | **0.36 (0.22 - 0.60)** | **0.48 (0.30 - 0.77)** |
| **Educational status** |  |  |
| No education | **Ref (1.00)** | **Ref (1.00)** |
| Primary | **1.42 (1.05 - 1.94)** | - |
| Secondary | **2.71 (2.03 - 3.58)** | **1.41 (1.14 - 1.75)** |
| Tertiary | **3.13 (1.65 - 5.94)** | - |
| **Wealth index** |  |  |
| Poorest | **Ref (1.00)** | **Ref (1.00)** |
| Poorer | **1.83 (1.26 - 2.66)** | - |
| Middle | **2.66 (1.82 - 3.90)** | - |
| Richer | **3.77 (2.42 - 5.86)** | **1.51 (1.07 - 2.12)** |
| Richest | **4.36 (2.64 - 7.21)** | **1.71 (1.45 - 2.54)** |
| **Frequency of listening to radio** |  |  |
| Not at all | **Ref (1.00)** | **Ref (1.00)** |
| < once a week | **2.14 (1.56 - 2.94)** | - |
| ≥ once a week | **2.77 (2.04 - 3.74)** | **1.37 (1.12 - 1.69)** |
| **Frequency of watching television** |  |  |
| Not at all | **Ref (1.00)** | **Ref (1.00)** |
| < once a week | **2.91 (2.05 - 4.14)** | - |
| ≥ once a week | **2.48 (1.85 - 3.34)** | - |
| **Place of residence** |  |  |
| Urban | **Ref (1.00)** | **Ref (1.00)** |
| Rural | **0.46 (0.34 - 0.62)** | **0.74 (0.61 - 0.92)** |
| *Continued* |  |  |

**Table 3 continued:** Logistic regression model for ANC determinants
| **Variables** | **COR (95%CI)** | **AOR (95%CI)** |
| --- | --- | --- |
| **Region** |  |  |
| Western | **Ref (1.00)** | **Ref (1.00)** |
| Upper West | **3.03 (1.19 - 7.73)** | **4.13 (2.22 - 7.68)** |
| Volta | **2.70 (1.04 - 7.02)** | **3.20 (1.55 - 6.58)** |
| Bono | **1.98 (1.73 - 5.39)** | **2.71 (1.32 - 5.56)** |
| Ahafo | **1.87 (1.78 - 4.49)** | **2.91 (1.52 - 5.57)** |
| Greater Accra | 1.71 (0.70 - 4.13) | - |
| Eastern | **1.65 (1.26 - 4.37)** | **2.15 (1.12 - 4.11)** |
| Oti | **1.25 (1.05 - 2.91)** | **2.57 (1.53 - 4.31)** |
| Ashanti | 1.20 (0.54 - 2.66) | - |
| Central | 1.19 (0.50 - 2.82) | - |
| Bono East | 0.57 (0.24 - 1.34) | - |
| Upper East | 0.55 (0.23 - 1.29) | - |
| Western North | 0.50 (0.23 - 1.03) | - |
| Northeast | **0.32 (0.16 - 0.64)** | **0.62 (.47 - 0.83)** |
| Northern | **0.27 (0.13 - 0.55)** | **0.59 (0.45 - 0.78)** |
| Savannah | **0.26 (0.13 - 0.54)** | **0.50 (0.38 - 0.66)** |
| **Women empowerment** |  |  |
| **Attitude towards violence** |  |  |
| Low empowerment | **Ref (1.00)** | **Ref (1.00)** |
| Medium empowerment | 1.29 (0.87 - 1.92) | - |
| High empowerment | **2.97 (2.15 - 4.11)** | **1.92 (1.53 - 3.10)** |
| **Social independence** |  |  |
| Low empowerment | **Ref (1.00)** | **Ref (1.00)** |
| Medium empowerment | 1.06 (0.80 - 1.41) | - |
| High empowerment | **2.26 (1.67 - 3.06)** | **1.82 (1.69 - 1.99)** |
| **Decision taking** |  |  |
| Low empowerment | **Ref (1.00)** | **Ref (1.00)** |
| Medium empowerment | **1.53 (1.08 - 2.17)** | - |
| High empowerment | **2.32(1.64 - 3.28)** | **1.31 (1.09 - 1.58)** |
| COR: crude odds ratio, AOR: adjusted odds ratio; Data in bold are significant at p-value < 0.05 |  |  |

Similarly, compared with those in the poorest wealth index, being in the Poorer wealth index (COR: 1.83; 95% CI: 1.26-2.66), middle wealth index (COR: 2.66; 95% CI: 1.82-3.90), richer wealth index (COR: 3.77; 95% CI: 2.42-5.86), and richest wealth index (COR: 4.36; 95% CI: 2.64-7.21) were more likely to receive quality antenatal care. Compared with pregnant women with lower empowerment levels, those with higher empowerment levels in the ‘attitude toward violence’ (COR: 2.97; 95% CI: 2.15 - 4.11), ‘social independence’ (COR: 2.26; 95% CI: 1.67 - 3.06) and ‘decision making’ (COR: 2.32; 95% CI: 1.64 - 3.28) domains were more likely to receive quality antennal care.

On the contrary, place of residence (rural or urban) and region of residence were associated with a decreased likelihood of receiving quality antenatal care. For example, women in rural areas were 54% less likely to receive quality antenatal care compared with pregnant women dwelling in urban areas (COR: 0.46; 95% CI: 0.34 - 0.62).

In the multivariable logistics regression model (**Table 3**), maternal age between 35-49 years, religion (other and no religion), pregnant women with secondary education, being in the richer and richest wealth index, listening to a radio at least once a week, region of residence (Upper West, Volta, Bono, Ahofo, Eastern, and Oti), pregnant women with higher empowerment in the ‘social independence’ and ‘decision making’ domains remained independently associated with increase in quality antenatal care. On the other hand, religion (other and No religion), living in rural area, region of residence (Northeast, Northern and Savannah) were independently associated with decrease in quality antenatal care.

For example, increase in maternal age (45-49 years), being in the richest wealth index, listening to a radio at least once a week, having higher empowerments in the ‘social independence’ and ‘decision’ making were independently associated with 5 times (AOR: 4.55; 95% CI: 2.76 - 7.49), 2 times (AOR: 1.71; 95% CI: 1.45 - 2.54), 1.4 times (AOR: 1.37; 95% CI: 1.12 - 1.69), 2 times (AOR: 1.82; 95% CI: 1.69 - 1.99), and 1.3 times (AOR: 1.31; 95% CI: 1.09 - 1.58) more likely to receive quality antenatal care, respectively. Interestingly, pregnant women residing in the Upper West, Volta, Bono, and Ahofo regions were 4 times, 3 times, 2.7 times and 2.9 times more likely to receive quality antenatal care, respectively. Contrarily, pregnant in rural regions such as the Northeast, Northern and Savannah regions were 38%, 41%, and 50% less likely to receive quality antenatal care, respectively.

## Discussion

In this study, we examined the association between sociodemographic and women empowerment variables with quality antenatal care in the 2022 GDHS. We found that women’s empowerment (“attitude toward violence”, “social independence”, and “decision-making”) domains, higher education, higher wealth index, older age, and media exposure were positively associated with quality ANC in Ghana with rural/urban and geographical disparities.

Similar to our findings, recent peer-reviewed studies have consistently demonstrated a positive association between women’s empowerment and increased utilization of antenatal care (ANC) services. For instance, greater women’s empowerment is associated with more antenatal visits regardless of economic status[22], higher quality antenatal services as women become more proactive in seeking comprehensive pregnancy care[23, 24] with economic empowerment having an added advantage[23]. Measures integrating women’s empowerment interventions into maternal and child health programs such as community-based education programs that enhance women’s healthcare decision-making autonomy, using the SWPER to monitor the effectiveness of women empowerment programs, enforcing laws against gender-based violence that hinder women’s healthcare-seeking behaviours, and policies on equal access to economic opportunities for women, will increase access to quality ANC.

Our findings indicated that higher maternal education, higher wealth index, older age, and media exposure (radio/TV) are associated with the likelihood of receiving quality ANC. The findings of analyses of 3-year (2018-2020) DHSs from 10 sub-Saharan countries also highlighted that women with higher educational levels and those in the higher wealth category have higher odds of adequate ANC compared to those with no formal education and lower wealth category[23]. In addition, a similar pooled study of 37 Low-and-Middle-Income countries also supported the evidence that older age >24 years and mothers’ exposure to media was positively associated with higher odds of ANC utilization[15]. Measures such as the promotion of girls’ education, especially in rural areas, expanding women’s economic empowerment initiatives, using the media to promote ANC awareness, and engaging religious and community leaders in maternal and child health advocacy will enhance quality ANC.

We observed geographical and rural versus urban disparities in quality ANC in Ghana. Similar findings indicated there were regional disparities in ANC attendance, with lower attendance in the Northern, North East, Savannah and Oti regions of Ghana[9].Findings from a cohort study in Vietnam also supported the rural/urban disparities in ANC, where women in urban were 5.2 times more likely to have adequate ANC compared to rural women[25]. Evidence from 27 Sub-Sharan countries added that the rural-urban disparity in maternal healthcare utilisation is mainly due to household wealth index, media exposure, and women’s educational level[26]. This implies women’s socio-economic empowerment initiatives, in addition to the expansion of healthcare infrastructure in rural and underserved areas and incentivizing healthcare providers to work in rural areas, will ultimately bridge the rural/urban quality ANC gaps. The stronger effect of empowerment in urban settings may reflect greater healthcare accessibility, better media exposure and more progressive gender norms. Future interventions should focus on increasing health infrastructure and advocacy in rural areas.

There are potential limitations in our study including situations where participants may not accurately recall ANC visits, and some regions may have been underrepresented in the dataset. Women with higher empowerment levels may have greater healthcare access, education, and economic stability, potentially introducing self-selection bias leading to overestimation of the true effect of empowerment on quality ANC. To address this, we conducted a Heckman two-step selection model. The coefficient on the IMR was not statistically significant (β = -0.08, p= 0.72), suggesting that selection bias is unlikely to affect our results. However, our findings should be interpreted with these possible limitations in mind. We recommend future longitudinal analyses to determine if consistent women’s empowerment results in sustained improvements in maternal health outcomes.

## Conclusion

In conclusion, our study provides strong evidence that empowering women, expanding educational opportunities, leveraging media for ANC awareness, addressing regional disparities and aligning ANC care with the World Health Organisation recommendations can significantly improve the quality of ANC. Achieving Ghana’s maternal health targets under the Sustainable Development Goals 3 (Good Health and Well-being) and 5 (Gender Equality) will require policies that enhance women’s decision-making power, economic autonomy and healthcare access.

## Data Availability

Data is available at DHS website upon request

## Abbreviations

ANC: Antenatal Care
AIC: Akaike Information Criterion
AOR: Adjusted Odds Ratio
COR: Crude Odds Ratio
DHS: Demographic and Health Survey
GDHS: Ghana Demographic and Health Survey
ICF: Inner City Fund
IMR: Inverse Mills Ratio
LMICs: Low- and Middle-Income Countries
SDGs: Sustainable Development Goals
SWPER: Survey-based Women’s empowerment
USAID: United States Agency for International Development
VIF: Variance Inflation Factor
WHO: World Health Organization

## Acknowledgements

We express our profound gratitude to the DHS Program for granting access to the dataset.

## Authors’ contributions

BB contributed to conceptualization, data analysis, interpretation of results, drafting of manuscript, and critical review and revision. MHK: contributed to conceptualization, data acquisition and analyses, interpretation of results and critical review and revision of manuscript. LB: contributed to conceptualization, data acquisition, draft of manuscript, critical review and revision of the manuscript. All authors reviewed the manuscript, contributed to the methodological framework, provided substantial intellectual input, and approved the final version.

## Funding

No funding was received for this study.

## Declarations

## Ethics approval and consent to participate

Not applicable.

## Consent for publication

Not applicable.

## Availability of data and materials

Data used for this analysis is available to the public upon request at https://dhsprogram.com/

## Competing interests

The authors declare that they have no competing interests.

## Notes

### Competing Interest Statement

The authors have declared no competing interest.

### Funding Statement

No funding for this work

